# In utero HIV exposure and anthropometry trajectories from birth through 8 years of age: findings from a prospective birth cohort in South Africa

**DOI:** 10.64898/2025.12.03.25341573

**Authors:** Angela M. Bengtson, Jennifer Pellowski, Maresa Botha, Tiffany Burd, Lesley Workman, Elizabeth Goddard, Dan J Stein, David Burgner, Toby Mansell, Heather J. Zar

## Abstract

**Background:** Children who are HIV-exposed but uninfected (CHEU) may have suboptimal growth, but few data are available beyond infancy to inform public health strategies.

**Methods:** We investigated anthropometry trajectories from 6 weeks to 8 years in a South African birth cohort, the Drakenstein Child Health study. Anthropometry was assessed at least annually by trained study staff and converted to weight-for-age (WAZ), height-for-age (HAZ), and body mass index (BMIZ) z-scores. Stunting (HAZ <-2SD from 12 months) and overweight (BMIZ score >2 SD from 6 months) were secondary outcomes. Multivariable linear mixed effects models were used to estimate associations between HIV exposure status and anthropometry trajectories and explore the impact of maternal HIV factors among CHEU.

**Findings:** Among 1,072 children (CHEU n= 236 (22%), children unexposed to HIV (CHU) n= 836 (78%)) mean birthweight was 3035 grams(g) (SD 592); CHEU 3012g (SD 598) vs CHU 3041g (SD 590) and 15.7% of infants were preterm (18.3% CHEU vs 15.0% CHU). Among women with HIV, 99% were on antiretroviral therapy (ART; 80% efavirenz-based ART), and 65% had an undetectable viral load in pregnancy. In multivariable analyses, CHEU had lower WAZ (marginal difference (MD) −0.16 (95% CI −0.32, −0.01) and HAZ (MD −0.26, 95% CI −0.41, - 0.11) scores, compared to CHU. Differences were largest before 3 years, but similar between CHEU and CHU thereafter. There was no association between HIV exposure and BMIZ scores (MD −0.02, 95% CI −0.17, 0.12), stunting (MD 0.05, 95% CI −0.04, 0.13) or overweight (MD 0.05, 95% CI −0.04, 0.13). Among CHEU, timing of maternal ART initiation, ART regimen, and viral load in pregnancy were not associated with anthropometry outcomes.

**Interpretation:** Compared to CHU, CHEU had lower weight and height from birth to 8 years, driven by early life differences. Among CHEU, maternal HIV factors did not drive anthropometry outcomes.

## Introduction

The global scale-up of antiretroviral therapy (ART) for people living with HIV has led to an increase in children exposed to HIV *in utero*, but uninfected (CHEU). Globally, there are over 14 million CHEU; nearly a quarter of whom live in South Africa(1), which has the highest burden of HIV in the world. Despite being born HIV-uninfected, CHEU may be more likely to experience suboptimal growth in infancy, compared to children unexposed to HIV (CHU), even in the era of universal ART.(2–4) Impaired growth in early life is associated with worse developmental, educational, economic, and health outcomes in childhood and later life.(5, 6) Improving growth outcomes for CHEU has been identified as a key public health priority to ensure CHEU are not just living HIV-free, but thriving.(7) However, there are few data available beyond infancy(8–10) or from low- and middle-income countries, where the burden of CHEU is highest (11), to inform public health strategies to optimize growth.

The mechanisms underlying suboptimal growth in CHEU are unclear, but likely multifactorial involving maternal, biologic, behavioral, social and structural factors. CHEU infants may be more likely to be born smaller(9), breastfed for shorter duration(12) and to experience a higher burden of infections in early life(13–16), all of which may affect growth. Among CHEU, maternal immunologic status, including viral load suppression(17) and timing and type of ART (18–20) may also influence growth, although findings have been mixed. Growth patterns in CHEU may also differ by sex and differences with CHU may become more pronounced during puberty. (21) Critically, whether deficits in growth among CHEU in early life persist into later childhood and have clinically meaningful implications for child health and development, remains unclear.(7) To better understand the drivers and long-term impacts of in utero HIV exposure on growth, longitudinal data on comparable groups of CHEU and CHU who are followed from birth into childhood are needed.

To address this gap, we examine anthropometry trajectories in a cohort of CHEU and CHU in a birth cohort from the same communities in South Africa. We examine differences in anthropometry trajectories from birth through 8 years of age by HIV exposure status, potential differences by breastfeeding status and sex, and among CHEU, explore potential differences in anthropometry trajectories by maternal HIV and ART factors.

## Methods

### Study population and design

We investigated anthropometry trajectories over time in children enrolled in the Drakenstein Child Health Study (DCHS), a longitudinal birth cohort of CHEU and CHU children from the same peri-urban communities outside of Cape Town, South Africa.(22) The study communities include a stable population (e.g. little in- or out-migration) of ∼200,000 and are characterized by high rates of poverty, substance use, and HIV infection.(23) Study communities have a well-established public healthcare system, which includes primary care, antenatal, obstetric, HIV, and prevention of mother-to-child-transmission services available free of charge in accordance with national guidelines.(24)

Details of the DCHS have been published previously.(22) Pregnant women were recruited from two primary health care clinics, Mbekweni (serving a predominantly black African community) and TC Newman (serving a mixed ancestry community). Mothers were enrolled in the DCHS while attending routine antenatal care and were prospectively followed. Women were eligible for the study if they were 18 years or older, between 20–28 weeks gestation, planned attendance at one of the two recruitment clinics and intended to remain in the area.(22, 25) Between 5 March 2012 and 31 March 2015, 1225 pregnant women were enrolled; 88 (7.2%) mothers were lost to follow up antenatally, had a miscarriage or a stillbirth. In total, 1137 women gave birth to 1143 live infants (4 sets of twins and 1 set of triplets) from 29 May 2012 to 3 September 2015.

Women with HIV enrolled into the DCHS received ART during pregnancy in accordance with South African HIV guidelines at the time.(26–29) Women and their children were followed through delivery and at least annually thereafter.(22) Of the 1143 live births, we excluded from the present analysis 5 infants who died prior to discharge from the hospital, 2 HIV exposed children who seroconverted during follow-up, and 64 children without anthropometry data after birth.

The DCHS was approved by the Faculty of Health Sciences, Human Research Ethics Committee, University of Cape Town (401/2009) and by the Western Cape Provincial Health Research committee. Mothers provided informed consent at enrolment and were re-consented annually. Consent was done in mother’s preferred language: English, Afrikaans or isiXhosa.

Assent was obtained annually from children from 7 years of age depending on their neurocognitive ability.

### Measures

The exposure of interest was HIV-exposure status during gestation for children. Women with unknown HIV status at study enrollment during pregnancy underwent HIV testing as part of routine care.(24) Women who tested negative for HIV at study enrollment were re-tested for HIV in the third trimester when feasible, and approximately every 3 months while breastfeeding as part of routine clinical care. HIV-exposed infants were given nevirapine syrup prophylaxis and tested for HIV at 6 weeks, 6-12 months, and 18 months of age, as part of routine care.(24)

We investigated anthropometry trajectories over time. Anthropometry, including length or height and weight, was assessed at least annually at all study visits by trained study staff using standardized protocols and regularly calibrated equipment.(30) Continuous measures were converted to age- and sex-adjusted weight for age (WAZ), height for age (HAZ), and body mass index (BMIZ) z-scores based on WHO child growth standards for term births (≥37 weeks).(31) Outliers for anthropometry z-scores were identified (typically <-6SD,>6SD) and recoded as missing according to WHO recommendations(32). WAZ and HAZ measures were adjusted for infants born preterm (<37 weeks gestation) through 50 weeks of age using the Fenton standards.(33, 34) BMIZ scores are not defined for preterm infants and were excluded prior to 50 weeks of age. (33, 34) We evaluated binary measures of stunting (HAZ <-2SD) from 12 months of age and overweight (BMIZ score >2 SD) from 6 months of age(32), when these outcomes are more likely to become clinically apparent. Changes in stunting and overweight status were descriptively evaluated over time.

Covariates of interest included maternal factors measured at 20-28 weeks of pregnancy, including age, BMI, educational attainment, parental employment, composite score of asset ownership, and monthly household income (in South African Rand), perinatal depression, intimate partner violence, and substance use.(35) Perinatal depression was evaluated using the Edinburgh Postnatal Depression Scale and a score of ≥13 (36) was used to define depressive symptoms. Intimate partner violence assessment was adapted from the WHO multicounty study(37) for use in Africa(38) and was defined as any report of sexual, physical, or emotional violence in the past 12 months. Maternal self-reported tobacco and alcohol use during the three months prior to the assessment was assessed using the WHO’s Alcohol, Smoking and Substance Involvement Screening Test (ASSIST)(39) and dichotomized into “any use” or “no use”. Delivery and postnatal covariates included: birth weight and length z-scores(32) and breastfeeding status. Breastfeeding status was defined as exclusive breastfeeding for at least one month versus not, based on the distribution of exclusive breastfeeding in the cohort.

Information on pregnancy complications, including pre-eclampsia, eclampsia, hypertension in pregnancy, and gestational diabetes were collected by study staff during pregnancy and via chart review. However, prevalence of pregnancy complications was low (<5%), thus we did not consider them in this analysis.(25) Study data were collected and managed using REDCap.(40, 41)

### Statistical Analysis

The goal of the statistical analysis was to evaluate associations of *in utero* HIV exposure status with anthropometry trajectories from 6 weeks until 8 years of age. We used linear mixed effects models with random effects to account for clustering by child, to estimate average WAZ, HAZ, and BMIZ trajectories over time by HIV exposure status and used mixed effects Poisson models with a log link and robust variance estimator to estimate the probability of stunting or overweight by HIV exposure status over time. Because growth is nonlinear, all models included an interaction term for HIV exposure status and time to allow associations to vary over time by HIV exposure status and were adjusted for maternal factors during pregnancy and birth weight and length z-scores, identified using a directed acyclic graph (Figure S1).(42) Postnatal factors that may influence growth are likely on the causal pathway between HIV exposure status and anthropometry trajectories and therefore were not included as covariates. We descriptively evaluated changes in stunting and overweight status over time, overall and by HIV exposure status, using Sankey plots. We explored potential effect measure modification by infant sex and exclusive breastfeeding status in stratified analyses. We conducted sub-group analyses among CHEU of associations of timing of maternal ART initiation ( pre- versus post-conception), maternal ART regimen in pregnancy (monotherapy with zidovudine (AZT), efavirenz-based ART with one non-nucleoside reverse transcriptase inhibitor (NNRTI) and two nucleoside reverse transcriptase inhibitors (NRTI), or second line treatment with a protease inhibitor (PI)), and maternal viral load (undetectable <40 copies/mL vs detectable) in pregnancy with anthropometry trajectories. All analyses were conducted using Stata 15 (StataCorp Inc, College Station, Texas, USA).

## Results

We included 1,072 children (CHEU n= 236 (22%), CHU n= 836 (78%); Figure 1), followed from 6 weeks through 8 years of age, who contributed 16,735 visits (CHEU 21% and CHU 78%; Table S1). At cohort enrollment, average maternal age was 26 years (SD 5.7) and BMI was 28.3 (SD 2.0; Table 1). Overall, 62% of the cohort had lower than a secondary education, 50% were not currently working, and 52% of households made R1000-5000 (∼$90-450 USD at the time of the study) per month. Psychosocial and substance use issues were common with 24% of women reporting perinatal depression, 34% recent intimate partner violence, and 28% prenatal tobacco use. Among women with HIV, 59% initiated ART during the index pregnancy, 80% were on first line (at the time) efavirenz-based ART, 15% were on AZT monotherapy and 5% were on second line PI-containing ART, 65% had an undetectable viral load (out of 142 with viral load data) and 40% had a CD4 count ≥500 cells/mm^3^ (out of 188 with CD4 count data). At delivery, the mean birthweight was 3034g (SD 592); CHEU 3011g (SD 598) vs CHU 3041g (SD 590) and 15.7% of infants were preterm (18.3% CHEU vs 15.0% CHU). Exclusive breastfeeding for at least one month was less common among CHEU (35.2%), compared to CHU (61.7%).

**Figure 1.**
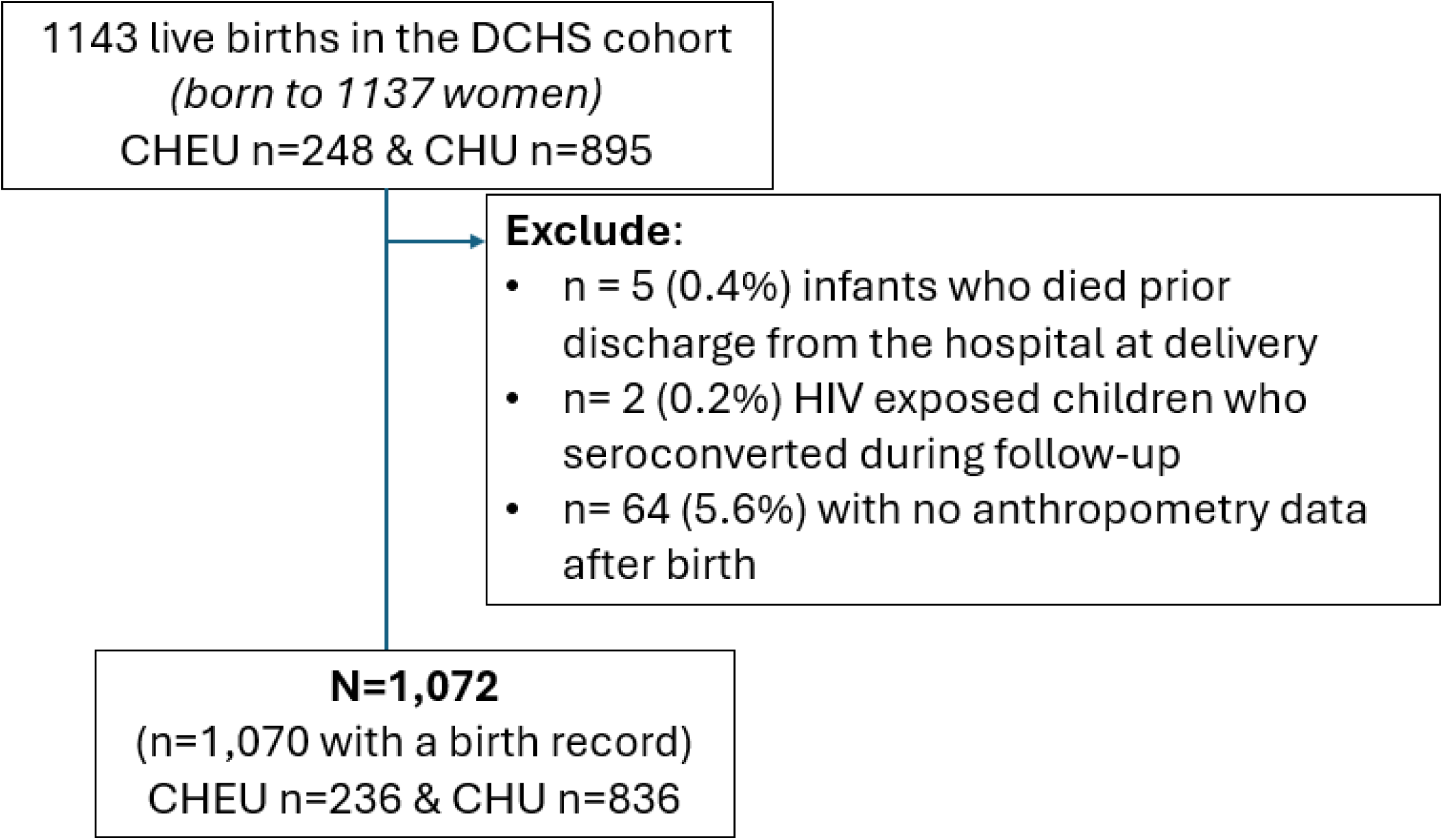
Study Population.

**Table 1.** Characteristics of participants in the Drakenstein Child Health Study.

|  | HIV exposed<br>uninfected<br>n=235 (22%) | HIV<br>unexposed<br>n=835<br>(78%) | Total<br>N=1070* |
| --- | --- | --- | --- |
| **Pregnancy characteristics (20-28 weeks gestation)** |  |  |  |
|  | Mean (SD) |  |  |
| Maternal age | 29.6 (5.2) | 25.8 (5.5) | 26.7 (5.7) |
| Maternal body mass index | 29.9 (6.2) | 27.8 (6.4) | 28.3 (2.0) |
| Asset summary score | 6.3 (2.0) | 7.0 (2.0) | 6.8 (2.0) |
|  | n (%) |  |  |
| Education |  |  |  |
| At least secondary or higher | 64 (27.4) | 347 (41.6) | 411 (38.48) |
| Lower than secondary | 170 (72.6) | 487 (58.4) | 657 (61.52) |
| Parental employment |  |  |  |
| Not working | 111 (47.2) | 420 (50.3) | 531 (49.63) |
| Working | 124 (52.8) | 415 (49.7) | 539 (50.37) |
| Household income (in Rand (R) per month) |  |  |  |
| <R1000 | 83 (35.3) | 288 (34.5) | 371 (34.71) |
| R1000-5000 | 125 (53.2) | 429 (51.4) | 554 (51.82) |
| >R5000 | 27 (11.5) | 117 (14.0) | 144 (13.47) |
| Perinatal depression<sup>1</sup> |  |  |  |
| No depressive symptoms | 153 (74.3) | 568 (76.0) | 721 (75.66) |
| Depressive symptoms | 53 (25.7) | 179 (24.0) | 232 (24.34) |
| Any recent intimate partner violence<sup>2</sup> |  |  |  |
| No | 148 (71.8) | 485 (64.9) | 633 (66.42) |
| Yes | 58 (28.2) | 262 (35.1) | 320 (33.58) |
| Maternal alcohol use<sup>3</sup> |  |  |  |
| No use | 203 (89.8) | 685 (86.3) | 888 (87.06) |
| Any use | 23 (10.2) | 109 (13.7) | 132 (12.94) |
| Maternal tobacco use<sup>3</sup> |  |  |  |
| No use | 206 (87.7) | 561 (67.2) | 767 (71.68) |
| Any use | 29 (12.3) | 274 (32.8) | 303 (28.32) |
| Site |  |  |  |
| Mbekweni | 219 (93.2) | 373 (44.7) | 592 (55.3) |
| TC Newman | 16 (6.8) | 462 (55.3) | 478 (44.7) |
| **Delivery and postnatal characteristics** |  |  |  |
|  | Mean (SD) |  |  |
| Gestational age at delivery (weeks) | 38 (2.6) | 38 (2.4) | 38 (2.4) |
| Birthweight (grams) | 3011 (598) | 3041 (590) | 3034 (592) |
| Birth weight z score | -0.3 (1.0) | -0.3 (1.1) | -0.3 (1.1) |
| Birth length z score | 0.3 (1.6) | 0.4 (1.6) | 0.4 (1.6) |
| - | n (%) |  |  |
| Infant sex |  |  |  |
| Female | 107 (45.5) | 416 (49.8) | 523 (48.9) |
| Male | 128 (54.5) | 419 (50.2) | 547 (51.1) |
| Preterm birth (delivery <37 weeks gestation) | 43 (18.3) | 125 (15.0) | 168 (15.7) |
| Exclusive Breastfeeding for >1 month | 82 (35.2) | 511 (61.7) | 593 (55.9) |

Missing data: \*n=2 infants missing information at birth (0.2%), education n=2 (0.2%), household income n=1 (0.1%), perinatal depression(n=117 (10.9%), intimate partner violence n=117 (10.9%), prenatal alcohol exposure n=50 (4.7%), maternal BMI n=33 (3.1%), birth length z score n=18 (1.7%). 1 Measured with the Edinburgh Depression Scale; $\geq 13$ indicates depressive symptoms. 2 Includes any physical, emotional or sexual violence within the last 12 months. 3 Based on self-report in the Alcohol, Smoking and Substance Involvement Screening Test (ASSIST).

Overall, mean WAZ scores were −0.27 (SD 1.18) at 6 weeks of age, increased to 0.18 (SD 1.29) at 9 months of age, and then declined to −0.31 (SD 1.28) at 8 years of age. BMIZ scores showed a similar trend to WAZ scores and were 0.15 (SD 1.14) at 6 weeks of age, increased to 0.88 (SD 1.58) by 18 months of age, and then declined to −0.17 (SD 1.24) at 8 years of age. Conversely, overall mean HAZ scores were −0.72 (SD 1.31) at 6 weeks of age, declined until 36 months to -1.14 (SD 1.14), and then increased but stayed below the WHO reference mean of 0 until 8 years of age (-0.35 (SD 1.05)) for CHU and CHEU combined.

In multivariable analyses of continuous z-scores, CHEU children had lower WAZ scores than CHU children overall (marginal difference −0.16; 95% CI −0.32, −0.01), which was driven by lower WAZ scores among CHEU in the first 9 months of life (mean difference range: −0.33 to - 0.19) with fewer differences by HIV exposure status after 12 months through 8 years of age (Figure 2). CHEU children also had overall lower HAZ scores (marginal difference −0.26, 95% CI −0.41, −0.11), with lower HAZ among CHEU through 3 years of age (mean difference range: - 0.43 to −0.16) before converging with CHU around 42 months of age and remaining similar through 8 years of age. There was no evidence of differences in BMIZ scores by HIV exposure status from 6 weeks through 8 years of age (marginal mean difference −0.02, 95% CI −0.17, 0.12).

**Figure 2.**
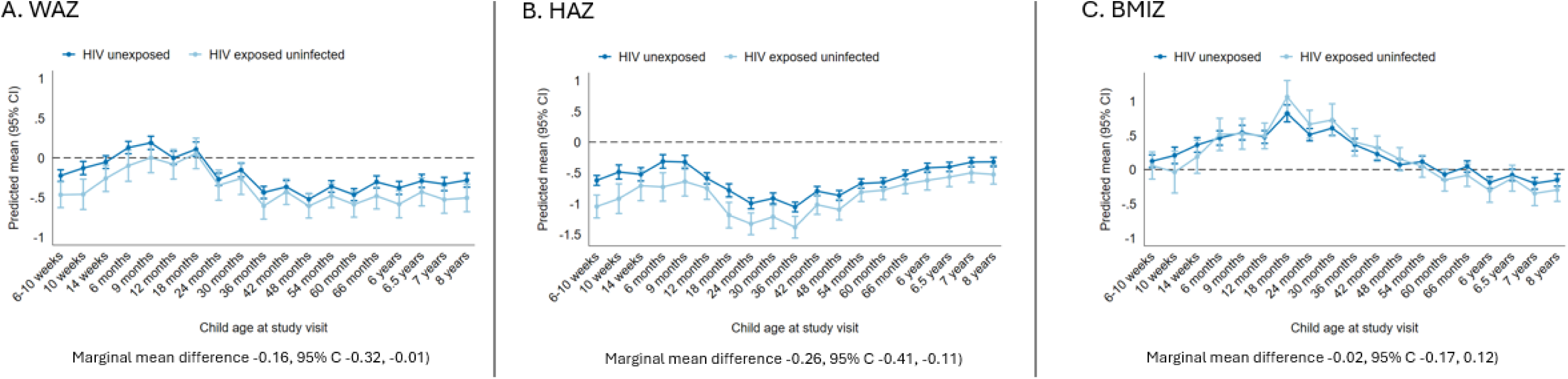
Weight for Age (WAZ), Height for Age (HAZ) and Body Mass Index (BMIZ) scores by HIV exposure status from 6 weeks to 8 years of age. Individual time points and marginal (overall) estimates from linear mixed effects models adjusted for clustering by child and for parental employment, household income, maternal education, household asset score, maternal BMI, maternal age, maternal depression, maternal IPV, maternal alcohol use, maternal smoking at cohort enrollment, birthweight zscore, birth length zscore, and site.

In the cohort the prevalence of stunting peaked at 3 years of age (22%) and then declined for both CHU and CHEU (Figure 3). In multivariable analyses, CHEU children had a higher probability of stunting through 3 years of age, however confidence intervals were wide and overlapping. There was no evidence that HIV exposure status was associated with stunting from birth through 8 years of age (marginal prevalence difference 0.05, 95% CI −0.04, 0.13). Most changes in stunting status occurred before age 5 years (Figure 4). The largest changes in stunting status were observed in children moving from being not stunted to stunted (range 6%-11%) between 12 and 36 months of age and in children who moved from being stunted at their previous visit to not stunted by 42 months of age (range 5%-10%), with few differences by HIV exposure status. After age 5, 4-7% of children remained stunted, 89-95% remained not stunted, and 0-3% either became stunted or moved from being stunted at their previous visit to being not stunted.

**Figure 3.**
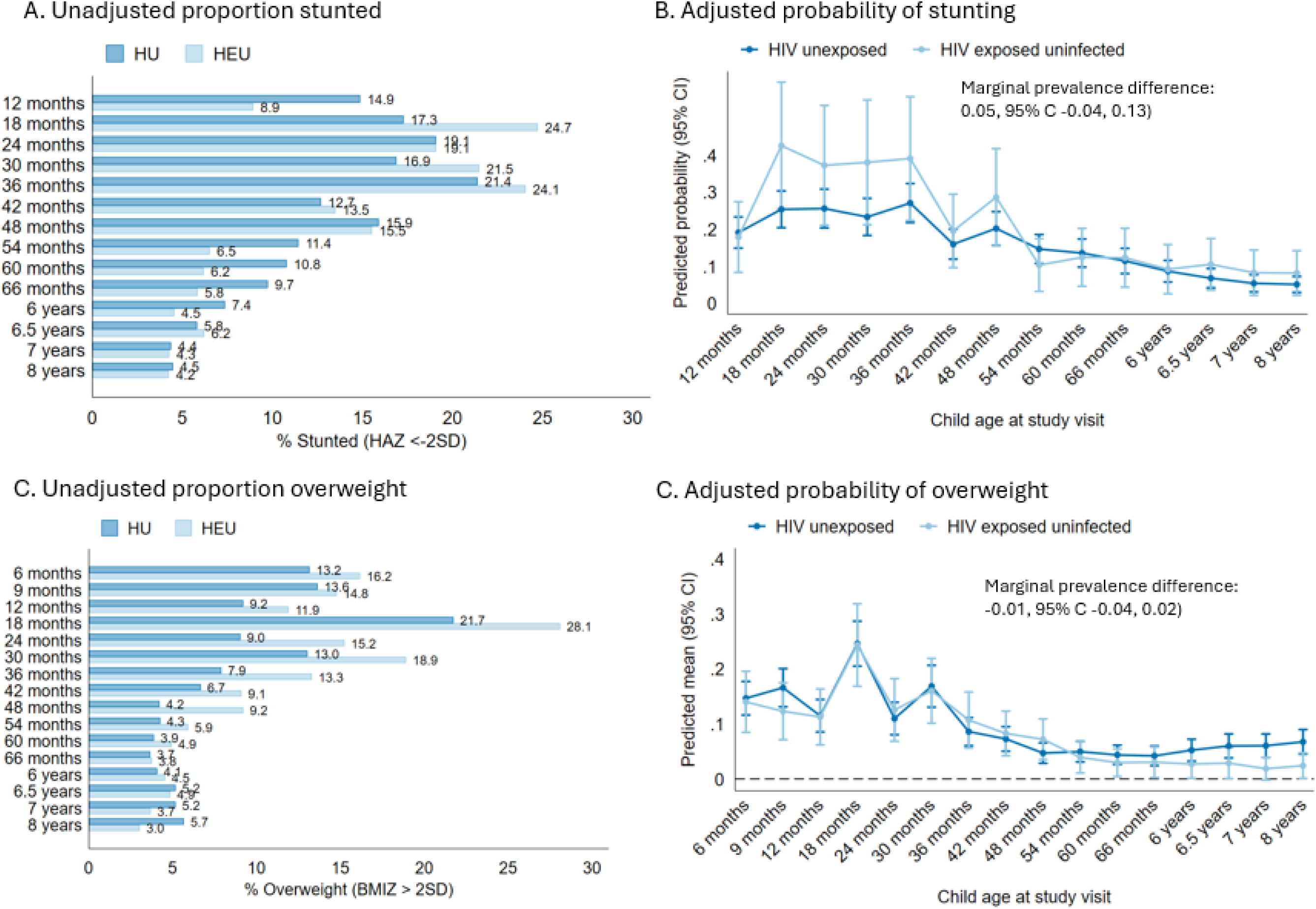
Probability of stunting (HAZ <-2 SD) and overweight (BMIZ > 2 SD) by HIV exposure status and age. Individual time points and marginal (overall) estimates from linear mixed effects models adjusted for clustering by child and for parental employment, household income, maternal education, household asset score, maternal BMI, maternal age, maternal depression, maternal IPV, maternal alcohol use, maternal smoking at cohort enrollment, birthweight zscore, birth length zscore, and site. HU=children HIV unexposed; HEU= children exposed to HIV but uninfected.

**Figure 4.**
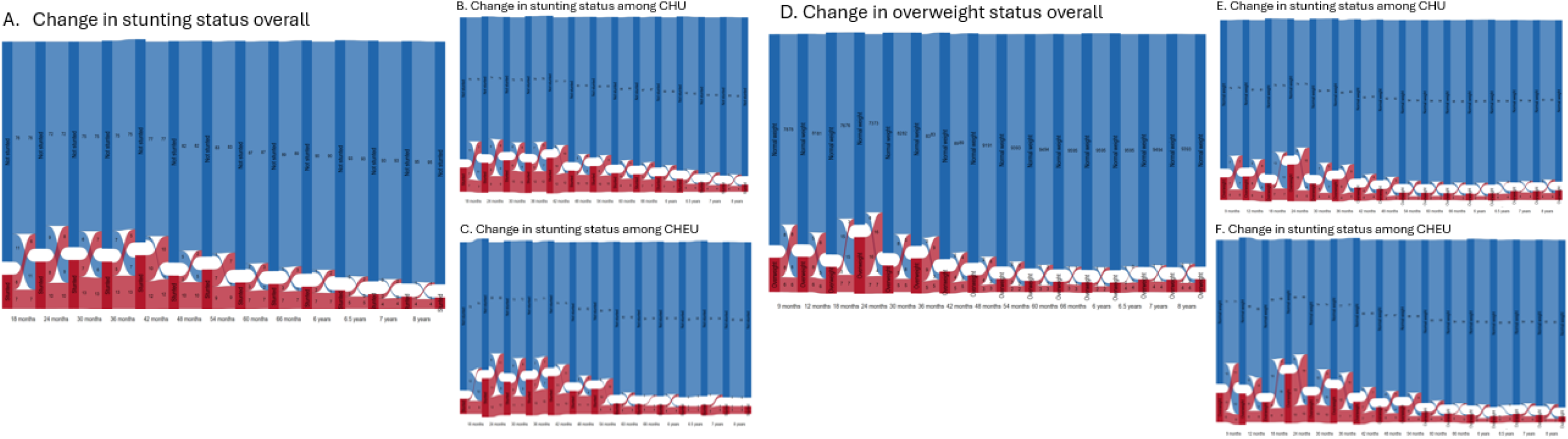
Change in stunting (HAZ < -2S0) and overweight (BMIZ > 2 SD) status by age and HIV exposure status.Numbers on the graphrepresent the proportion moving from onecategory to another category, defined by change in overweight or stuntingstatus, from their last study visit. Graphs are presented for the overall cohort (A, DJ, among children HIV-unexposed (CHU) children (B, E) and children HIV-exposedbut uninfected (CHEU) children (C, F). Blue indicates being not stunted (A-C) or overweight (D-E). Red indicates being stunted (A-CJor overweight (D-E).

The prevalence of being overweight was highest through 18 months of age (9-28%) before declining for both CHEU and CHU (Figure 3). In multivariable analyses, HIV exposure status was not associated with being overweight from 6 months through 8 years of age (marginal prevalence difference −0.01, 95% CI −0.04, 0.02). Changes in overweight status occurred predominately before age 3, with 3-15% of children moving from normal weight to overweight and 3-16% of children moving from overweight to not overweight between visits. Overall trends in change in overweight status were similar for CHEU and CHU, with a slightly higher proportion of CHEU remaining overweight (CHEU 6-10% versus CHU 4-7%) from 6 months through 3 years of age. After age 3 years, overweight status stabilized at 3-5% overweight, 90-95% not overweight and 1-3% moving from either not overweight to overweight or vice versa.

In exploratory multivariable stratified analyses evaluating sex and exclusive breastfeeding for at least 1 month as potential effect measure modifiers, differences in WAZ and HAZ scores by HIV exposure status were slightly larger among boys in early life, relative to girls (Figure S2).

Additionally, associations between HIV exposure status and HAZ were more pronounced among children not exclusively breastfed for at least a month, but similar among children exclusively breastfed at least 1 month irrespective of HIV exposure status (Figure S3). There was no evidence of effect measure modification by child sex or exclusive breastfeeding status for associations between HIV exposure status and BMIZ scores. In subgroup analyses restricted to CHEU children, there were few differences in WAZ, HAZ, or BMIZ scores by timing of maternal ART initiation, maternal ART regimen, or maternal viral load in pregnancy (Figure S4).

## Discussion

In this birth cohort of South African children, CHEU had lower height and weight z-scores through 8 years of age compared to CHU. These differences were driven by lower height and weight among CHEU within the first 1 to 3 years of life, with the largest differences seen among CHEU boys and those not exclusively breastfed for at least 1 month. Differences in weight and height between CHEU and CU were minimal after 3 years of age, when mean z-scores for weight and height were below the WHO reference mean irrespective of HIV exposure. Early life deficits in height and weight did not translate into an increased risk of stunting or being overweight for CHEU, compared to CHU, in the first 8 years of life. Among CHEU children with near universal ART exposure in pregnancy, there was little evidence of maternal HIV or ART factors influencing child growth from birth through 8 years of age.

Our finding that CHEU had impaired growth in early life, particularly related to length/height, but similar growth patterns to CHU after early childhood aligns with several other studies.(2, 3, 8, 43) A higher risk of stunting among CHEU has also been reported.(2, 4, 8) While we did not see differences in the risk of stunting in CHEU compared to CHU through 8 years of age, 22% of children were stunted at 3 years of age and CHEU had a higher prevalence of stunting within the first three years of life, compared to CHU. Notably, both CHEU and CHU children were approximately 0.5-1 standard deviation below the WHO reference mean for HAZ from 6 weeks through 8 years of life, indicating nutritional deficiencies in this population regardless of HIV exposure.(44)

The reasons for impaired growth among CHEU in early life are multifactorial but have been hypothesized to be related to being born lower birthweight (11). In this cohort, CHEU and CHU infants had similar mean weights at birth and similar proportions of preterm birth and marginal (overall) differences in HAZ and WAZ scores persisted after adjustment for birthweight and birth length z-score and other risk factors during pregnancy. While encouraging that CHEU ‘caught up’ to CHU in terms of height and weight in early childhood, growth faltering in early life is well documented to increase the risk of cardiometabolic disease later in life.(6) These findings suggest that CHEU may be at increased risk of cardiometabolic comorbidities in adulthood, relative to CHU, and that a clearer understanding of early life drivers of growth in CHEU, such as increased infection burden(14, 45), may also be required to optimize growth among CHEU.

In exploratory analyses, we observed smaller differences in weight and height between CHEU and CHU who were exclusively breastfed. Consistent with other South African studies(12), the duration of exclusive breastfeeding in this cohort was short and CHEU were less likely to be exclusively breastfed for at least a month. CHEU, who were exclusively breastfed for at least one month, had similar HAZ and WAZ scores as CHU within the first year of life, compared to CHEU children who were not exclusively breastfed for at least one month. While these findings are exploratory, breastfeeding has been associated with improvements in weight, height and development for both CHEU and CHU.(46) Public health efforts to support breastfeeding initiation and continuation may help to support the recommended 6 months of exclusive breastfeeding for CHEU and improve growth outcomes.

We also explored differences in early life growth by infant sex. Differences in weight and height by HIV exposure status were larger in boys than girls, as has been reported elsewhere.(17, 21) Sex-specific differences in fetal and infant growth within the general population are well documented.(47, 48) While boys tend to be larger on average at birth and in infancy in well-resourced settings, in the context of food insecurity there is increasing evidence that boys are at greater risk of undernutrition than girls.(49–51) The contribution of *in utero* HIV exposure to impaired growth among boys in resource-limited settings remains unclear, but is critical to understand in order to target resources to CHEU at highest risk of growth faltering.

We observed few differences in infant anthropometry by maternal HIV and ART factors in this cohort with high ART coverage and viral load suppression during pregnancy.(17, 19, 45) While we did not have measures of maternal ART adherence, poor ART adherence has been associated with stunting in CHEU.(52) The DCHS cohort was enrolled between 2012 and 2015, and therefore reflects in utero exposure to primarily efavirenz-based ART, which was first line treatment at the time.(27) First line HIV treatment in South Africa now includes dolutegravir, an integrase stand inhibitor which has been associated with weight gain in adults. Current evidence suggests that length/height may be improved in CHEU exposed to dolutegravir versus efavirenz (53), but that growth deficits relative to CHU remain in the context of dolutegravir exposure (45, 54).

Our analysis has strengths and limitations. Strengths include the ability to examine longitudinal anthropometry trajectories in a cohort of CHEU and CHU from the same community with little loss to follow up from infancy into childhood, with adjustment for a broad range of antenatal and birth variables. We also were able to explore potential postnatal modifiers of growth and, among CHEU, whether maternal HIV and ART drivers influenced anthropometry trajectories.

Limitations include the exclusion of BMIZ scores before 50 weeks gestation for infants born preterm, as they are not defined.

## Conclusion

In a South African cohort followed from birth through 8 years of age we observed small deficits in weight and height, but not body mass index, stunting or being overweight, among CHEU. These deficits were driven by lower WAZ and HAZ scores among CHEU, compared to CHU in early life. By 1-3 years of age CHEU and CHU displayed similar anthropometry trajectories.

Through 8 years of age, mean z scores for weight and height fell below the WHO reference mean for both CHEU and CHU, indicating that public health interventions to optimize growth for all children within this setting remain critical. While it is encouraging that CHEU had similar anthropometry trajectories to CHU beyond early childhood, early life growth deficits remain predictors of adverse health outcomes in later life. Future work should focus on understanding the drivers and clinical implications of early life growth deficits in early life among CHEU to optimize health outcomes.

## Data Availability

The Drakenstein Child Health Study is committed to the principle of data sharing. De-identified data will be made available to requesting researchers as appropriate. Requests for collaborations are welcome. More information can be found on our website [http://www.paediatrics.uct.ac.za/scah/dclhs].

## Acknowledgements

We thank the mothers and their children for participating in the study and the study staff, the clinical and administrative staff of the Western Cape Government Health Department at Paarl Hospital and at the clinics for support of the study.

## Target Journal

Lancet Child and Adolescent Health

## Supplemental Tables and Figures

**Figure S1.**
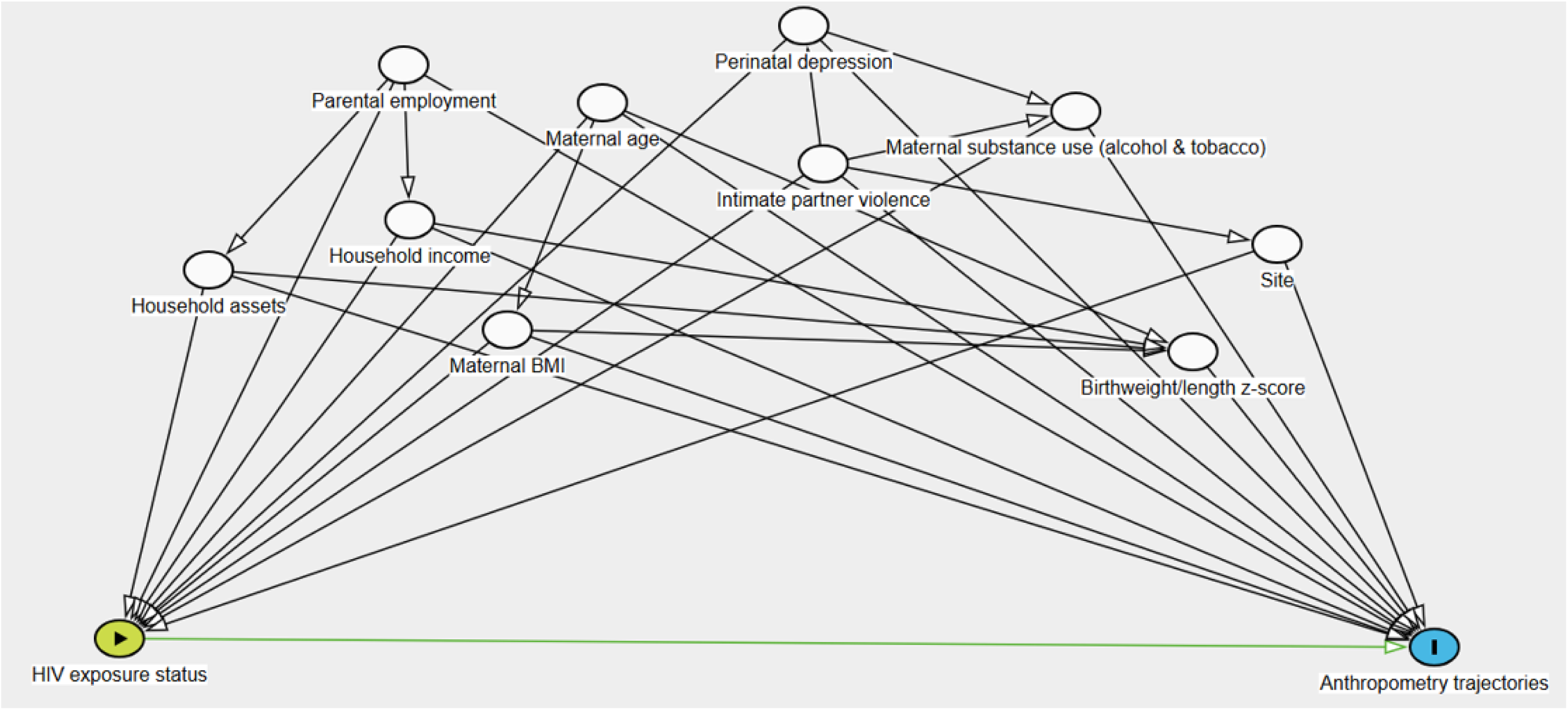
Directed Acyclic Graph. Except for birth weight and length z score, all covariates measured during pregnancy.

**Figure S2.**
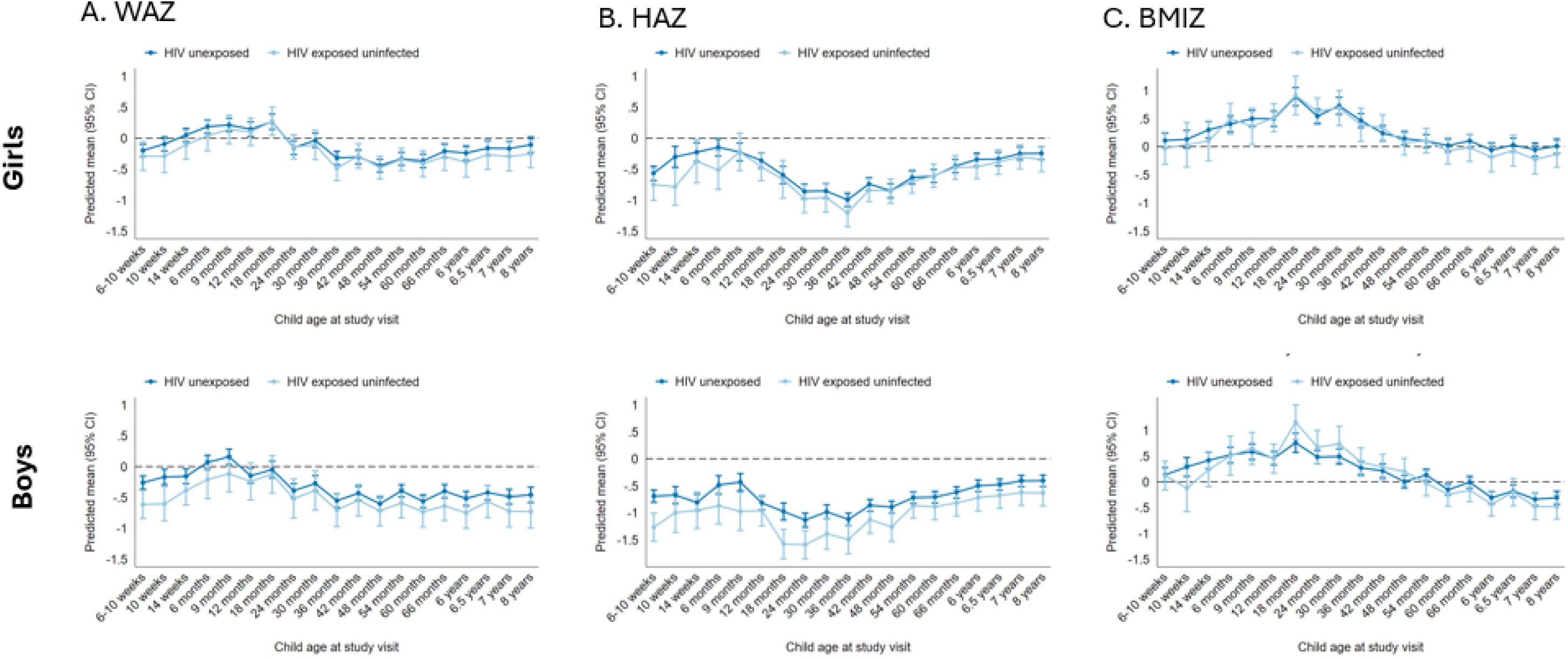
Weight for Age (WAZ), Height for Age (HAZ) and Body Mass Index (BMIZ) scores stratified by infant sex. Multivariable linear mixed effects models are adjusted for parental employment, household income, maternal education, household asset score, maternal BMI, maternal age, maternal depression, maternal IPV, maternal alcohol use, maternal smoking at cohort enrollment, birthweight zscore, birth length zscore, and site.

**Figure S3.**
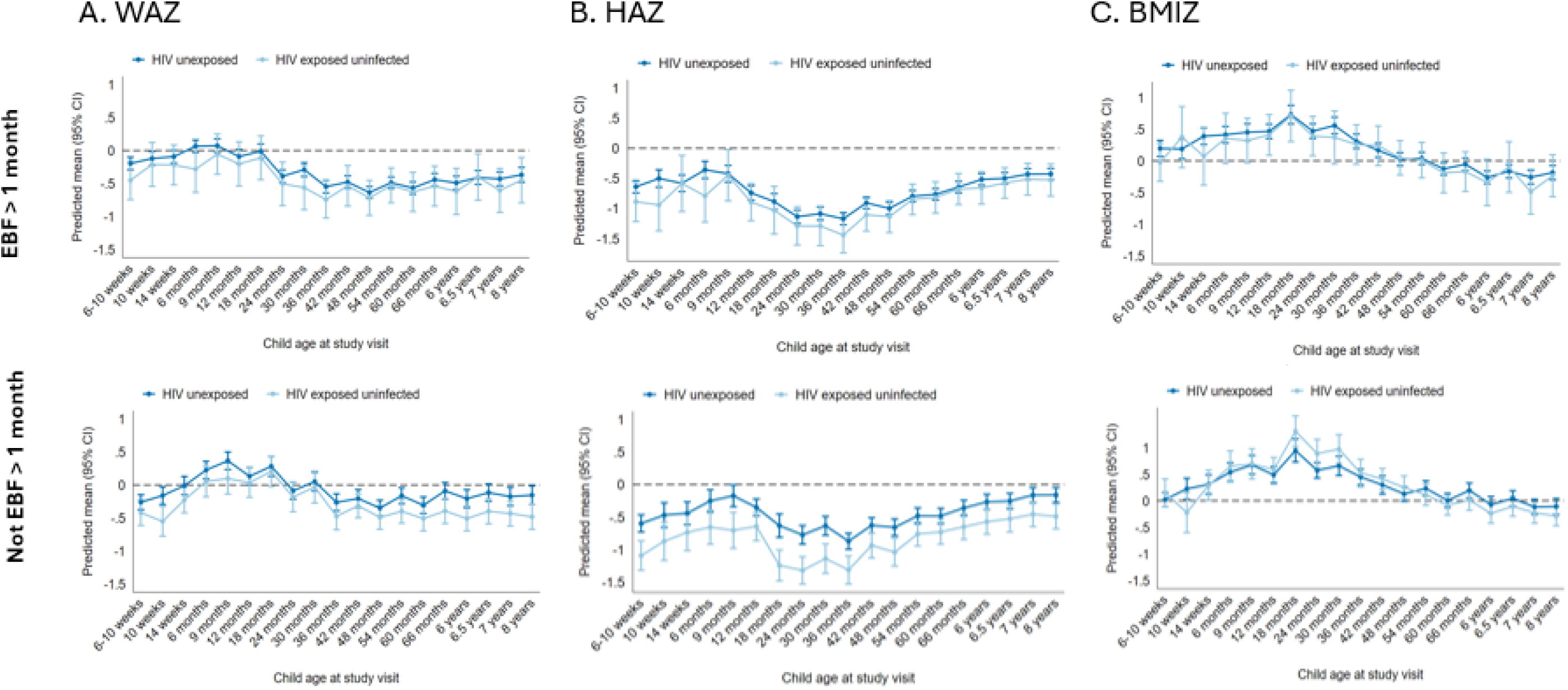
Weight for Age (WAZ), Height for Age (HAZ) and Body Mass Index (BMIZ) scores stratified by exclusive breastfeeding status (EBF). Multivariable linear mixed effects models are adjusted for parental employment, household income, maternal education, household asset score. maternalBMI, maternal age, maternal depression, maternal IPV, maternal alcohol use, maternal smoking at cohort enrollment, birthweight zscore, birth length zscore, and site.

**Figure S4.**
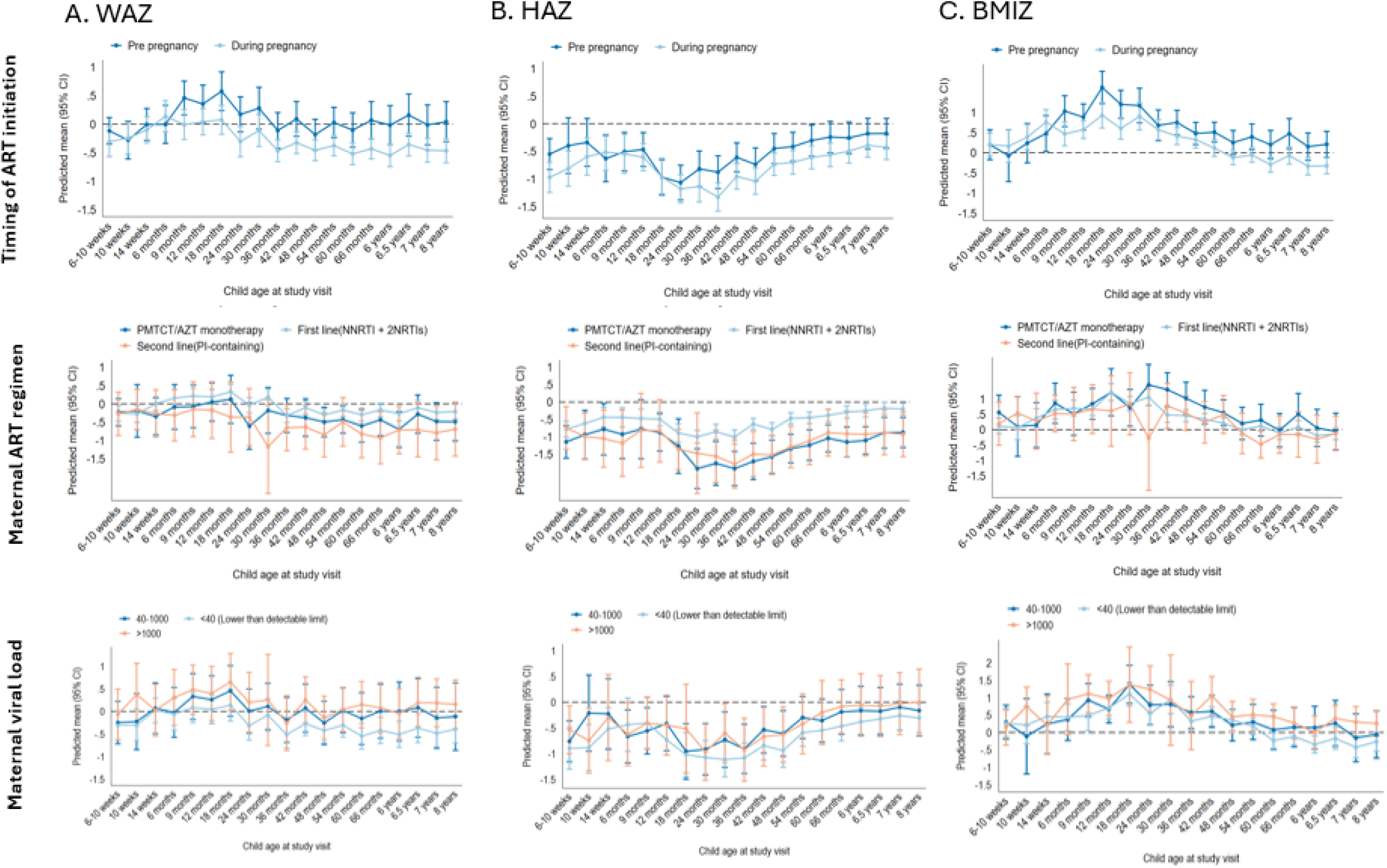
Subgroup analyses of Weight for Age (WAZ), Height for Age (HAZ) and Body Mass Index (BMIZ) scores among HIV exposed, but uninfected children. Multivariable linear mixed effects models areadjusted for parental employment, household income, maternal education, household asset score, maternal BMI. maternal age, maternal depression, maternal IPV, maternal alcohol use, maternal smoking at cohort enrollment, birthweight zscore, birth length zscore, and site.

**Table S1.**
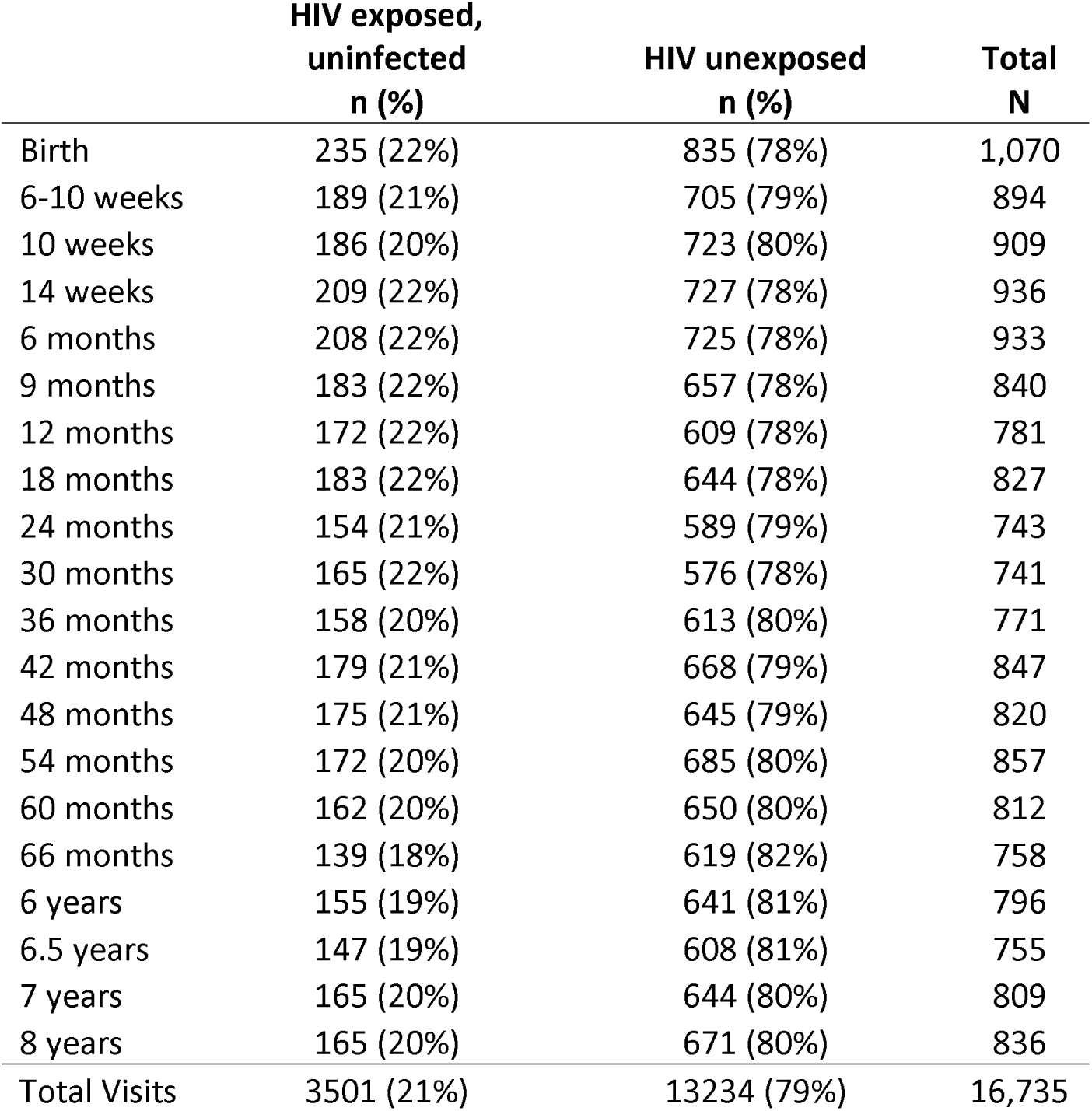
Study population at each study visit by child HIV exposure status.

|  | HIV exposed,<br>uninfected<br>n (%) | HIV unexposed<br>n (%) | Total<br>N |
| --- | --- | --- | --- |
| Birth | 235 (22%) | 835 (78%) | 1,070 |
| 6-10 weeks | 189 (21%) | 705 (79%) | 894 |
| 10 weeks | 186 (20%) | 723 (80%) | 909 |
| 14 weeks | 209 (22%) | 727 (78%) | 936 |
| 6 months | 208 (22%) | 725 (78%) | 933 |
| 9 months | 183 (22%) | 657 (78%) | 840 |
| 12 months | 172 (22%) | 609 (78%) | 781 |
| 18 months | 183 (22%) | 644 (78%) | 827 |
| 24 months | 154 (21%) | 589 (79%) | 743 |
| 30 months | 165 (22%) | 576 (78%) | 741 |
| 36 months | 158 (20%) | 613 (80%) | 771 |
| 42 months | 179 (21%) | 668 (79%) | 847 |
| 48 months | 175 (21%) | 645 (79%) | 820 |
| 54 months | 172 (20%) | 685 (80%) | 857 |
| 60 months | 162 (20%) | 650 (80%) | 812 |
| 66 months | 139 (18%) | 619 (82%) | 758 |
| 6 years | 155 (19%) | 641 (81%) | 796 |
| 6.5 years | 147 (19%) | 608 (81%) | 755 |
| 7 years | 165 (20%) | 644 (80%) | 809 |
| 8 years | 165 (20%) | 671 (80%) | 836 |
| Total Visits | 3501 (21%) | 13234 (79%) | 16,735 |

